# Herpes simplex virus type 1 DNA is less prevalent in persons with Alzheimer’s disease and genetic factors modify the effect

**DOI:** 10.64898/2026.04.16.26351043

**Authors:** Marlene Tejeda, John Farrell, Congcong Zhu, Lee Wetzler, Kathryn L. Lunetta, William S. Bush, Eden R. Martin, Li-San Wang, Gerard D. Schellenberg, Margaret A. Pericak-Vance, Jonathan L. Haines, Lindsay A. Farrer, Richard Sherva

**Affiliations:** Departments of Medicine Biomedical Genetics, Boston University Chobanian & Avedisian School of Medicine, Boston, MA, 02118, USA; Departments of Medicine Infectious Disease, Boston University Chobanian & Avedisian School of Medicine, Boston, MA, 02118, USA; Microbiology, Boston University Chobanian & Avedisian School of Medicine, Boston, MA, 02118, USA; Neurology, Boston University Chobanian & Avedisian School of Medicine, Boston, MA, 02118, USA; Ophthalmology, Boston University Chobanian & Avedisian School of Medicine, Boston, MA, 02118, USA; Departments of Biostatistics, Boston University School of Public Health, Boston, MA, 02118, USA; Departments of Epidemiology, Boston University School of Public Health, Boston, MA, 02118, USA; Department of Population & Quantitative Health Sciences, Cleveland Institute for Computational Biology, Case Western Reserve University School of Medicine, Cleveland, Ohio, 44106, USA; John P. Hussman Institute for Human Genomics and Dr. John T. MacDonald Foundation Department of Human Genetics, Miller School of Medicine, University of Miami, Miami, Florida, 33136, USA; Penn Neurodegeneration Genomics Center, Department of Pathology and Laboratory Medicine, University of Pennsylvania Perelman School of Medicine, Philadelphia, Pennsylvania, 19104, USA

**Keywords:** Alzheimer disease, herpes simplex virus type 1, whole-genome sequencing, host–virus interaction, APOE, genome-wide association study, viral DNA detection, neurodegeneration

## Abstract

**INTRODUCTION:** Herpes simplex virus-1 (HSV-1) has been implicated in Alzheimer disease (AD).

**METHODS:** Reads from Alzheimer’s Disease Sequencing Project whole-genome sequencing data collected from brain (2,203 AD; 616 controls) and blood (8,908 AD; 15,768 controls) were aligned to viral genomes. Generalized linear mixed-models tested for the effect of HSV-1 DNA on AD, and we performed GWAS on HSV-1 presence and SNP×HSV-1 interaction effects on AD, adjusting for age, sex, tissue, library preparation, relatedness, and ancestry principal components.

**RESULTS:** Across ancestry groups, HSV-1 DNA was consistently less frequent in AD cases; reads predominantly mapped to regions containing the latency-associated transcript region. DNA prevalence was lower in *APOE*-ε4 carriers; HSV-1 was associated with reduced AD risk in ε4 non-carriers but increased risk in carriers. GWAS identified host genetic influences on HSV-1 detection and interaction loci affecting AD risk.

**DISCUSSION:** HSV-1 DNA showed an inverse association with AD and is affected by genetics.

## 1. BACKGROUND

Alzheimer’s disease (AD) is a complex and chronic neurodegenerative condition that primarily affects the elderly. Despite substantial progress in understanding the fundamental mechanisms underlying AD, many aspects of the disease etiology remain enigmatic. Recent research has unveiled intriguing connections between AD and viral infections[1]. HSV-1 is a member of the α-herpesvirinae subfamily within the Herpesviridae family. Like other herpesviruses, HSV-1 undergoes distinct phases of infection, including latency and reactivation [2][3]. Although nearly everyone has been exposed to one or more herpes viruses worldwide, epidemiological studies indicate that approximately one-third of the population does not experience active or reactive infections[4]. These viruses are neurotropic and have been found in brain tissue from those with a pathological diagnosis of AD[5][6], potentially contributing to the deposition of toxic forms of amyloid-beta (Aβ) and Tau proteins, the hallmark pathological features of AD[5][7][8]. Production of Aβ increases in response to infection and may help protect the brain against infectious agents, including HSV-1[5]. HSV-1 infections also induce accumulation of Aβ_42_ inside neurons by a calcium-dependent mechanism [5][9].

We previously reported a significant, positive association of HSV-1 DNA detected in whole-exome sequence (WES) data derived from human blood and brain with AD risk [10]. These effects varied across ancestry groups, tissue source, and sequencing experiment type. In this study, we re-examined the viral DNA-AD association in a substantially larger sample of whole genome (WGS) data from multiple ancestry groups, and then tested for the effects of genetic variation on both the presence of viral DNA and as a modulator of viral infection in AD.

## 2. METHODS

### 2.1 Subject ascertainment and characteristics

WGS data were collected from blood and brain samples of participants in the Alzheimer’s Disease Sequencing Project (ADSP), which seeks to identify genetic factors influencing late-onset Alzheimer’s disease (AD) in multiple ancestral populations[11]. We obtained WGS and phenotypic information from the ADSP Release 4 (R4) dataset that includes individuals ascertained by ten Alzheimer Disease Research Centers and 17 other cohorts from 14 countries[12]. Some cohorts include families with multiple members affected with AD. The sample included 27,315 individuals with WGS data derived from brain tissue (N = 2,639) or blood samples (N = 24,676) (Table S1). Individuals with an unknown AD status (N=7,022) or tissue source (N=1,857) were excluded from further analyses. Cohort details and participant data are accessible through the National Institute on Aging Genetics of Alzheimer’s Disease Data Storage site (NIAGADS)[12][13].

### 2.2 Kinship and Population Substructure Analysis

We computed a KING kinship matrix using the GENESIS software [14] to specify biological relationships. Principal components (PCs) of ancestry were computed for unrelated individuals and projected onto related individuals using the PC-AIR method in GENESIS and reference populations from the 1000 Genomes Project and HGDP available in gnomAD. These global ancestry PCs were used to define ancestry groups and as covariates in GWAS analyses. Ancestry groups were derived by applying Gaussian Mixture models to multiple k-means cluster initializations based on the global PCs. This procedure assigns all individuals to one of the inferred groups, including admixed individuals, ensuring that no samples were left unclassified (Figure S1) . Assignment certainty was high, with the majority of individuals showing GMM membership probabilities above 0.90, and nearly all exceeding 0.50; only 16 admixed samples exhibited lower probabilities. European (EA; 5,464 AD cases, 3,517 controls), African American (AA; 1,527 AD cases, 3,017 controls), Indian (IND; 123 AD cases, 2,477 controls), native American Hispanic (NAH; 891 AD cases, 3,491 controls), and Caribbean Hispanic (CH; 2,014 AD cases, 2,799 controls), as well as several smaller clusters reflecting substructure within the European-ancestry group. These included an Ashkenazi Jewish cluster (AJ: 590 AD cases, 450 controls) and an admixed European–Middle Eastern (EA/MID: 818 AD cases, 485 controls) cluster (≈ 80% EUR and 20% MID), consistent with Mediterranean or Balkan ancestry. The AM cluster (AM: 94 AD cases, 598 Controls) closely overlaps with the EU cluster, both showing predominantly European ancestry and minimal admixture from other populations (Figure S1). Therefore, we combined AM with EU in downstream analyses. In contrast, the EU/MID and AJ showed Middle Eastern ancestry components.

Classifying individuals into ancestry groups was necessary both to provide ancestry-specific descriptive statistics and to enable subsequent within-ancestry kinship estimation using PCRelate. For mixed-model association, the Genetic Relationship Matrix (GRM) required by SAIGE was constructed using SAIGE’s sparse GRM approach, ensuring that population structure captured by the global PCs was appropriately incorporated. PCs were recomputed with KING within each ancestry group to obtain kinship estimates adjusted for population structure within ancestry. After identifying the unrelated samples within each group using PC-AIR, we selected a LD-pruned set of autosomal SNPs based on MAF ≥3%, missing <1%, sliding window 500,000 bp / 500 SNPs, and r² > 0.2. We calculated a second set of PCs from each pruned set and supplied the top 20 PCs to PCRelate (GENESIS) to generate a PC-adjusted kinship matrix that distinguishes true relatedness from residual population structure.

### 2.3 DNA sequence data processing and quality control

DNA sequencing, variant calling, QC procedures related to the sequencing data were previously reported [11,16,17]. Genotype data processing and cleaning were performed using PLINK 2.0 [18]. Quality control was conducted within each ancestry group, and single nucleotide polymorphisms (SNPs) with missing genotype rates >5%, significant deviation from Hardy-Weinberg equilibrium (p<0.05), and minor allele frequency (MAF) <1% were excluded. This threshold was chosen because it was within ancestry and effectively reduced mild genomic inflation in preliminary analyses.The total number of filtered variants by dataset and ancestry group is shown in Table S2.

### 2.4 Microbial DNA detection

To detect viral DNA within the human DNA sequence data, we developed MicrobeSeq, a custom pipeline and obtained complete reference genomes (FASTA files) for 318 viral species from the National Center for Biotechnology Information (NCBI)[19]. In brief, sequencing reads that mapped to the human genome (build GRCh38) were removed, resulting in a FASTQ file enriched with non-human DNA reads. FASTQ files were aligned to sequences from all microbe reference genomes using BWA-MEM [20]. Counts of viral reads that matched to 59 viral species were normalized to the original host alignment read depth. This initial screen showed that HSV-1 was detected in >50% of individuals across all ancestry groups, whereas the prevalence of other viruses identified in at least 50 samples was <4% and often limited to one or few groups (Table S3). These observations provided the rationale for focusing analyses on HSV-1. Reference coordinates for HSV-1 were obtained from the NCBI RefSeq genome NC_001806.2. The GenBank flat file was downloaded from NCBI (Entrez efetch) and programmatically parsed to extract genomic intervals annotated as repeat-long segments. The terminal repeat long (TRL) and internal repeat long (IRL) regions, which both encode the latency associated transcript (LAT) responsible for epigenetic repression of viral reactivation, were defined from the GenBank feature annotations and exported in BED format.

### 2.5 Statistical analysis methods

We used R software to implement a generalized linear model (GLM) to estimate the effect of HSV-1 viral presence on AD status within each ancestry group controlling for *APOE* ε2 and ε4 carrier status, sex, age, sequencing center, tissue source (blood vs. brain), and library preparation method (PCR-free vs. PCR-based). HSV-1 reads were present in both PCR-free and PCR-based samples, PCR-based WGS did not suppress viral detection. Library method was included as a covariate to adjust for any remaining bias.Genetic association analyses were limited to HSV-1 because of low power for tests of other viruses which are relatively infrequent. In addition, we tested whether the effect of HSV-1 on AD risk was modified by *APOE* genotype. We analyzed *APOE* separately from the genome-wide screen because *APOE* ε4 is the strongest known common genetic determinant of AD risk, and there is longstanding evidence that *APOE* genotype modifies susceptibility to HSV-1[21]. One GLM model evaluated AD status as the outcome with HSV-1 presence, *APOE* ε4 carrier status, and their interaction as predictors, and a second model tested HSV-1 presence as the outcome and *APOE* ε4 carrier and AD status as predictors, adjusting for the same covariates listed above. Both models were tested separately within each ancestry group.

To assess whether blood and brain samples differed in HSV-1 associations, we repeated the regressions within each tissue type. Brain-derived sample sizes within ancestry groups were too small to support ancestry-specific genome-wide association analyses (AA: N=131, AJ: N=316, CH: N=47, EA/MID: N=346, EA: N=1738, IND: N=0, NAH: N=61).

We extended these two models genome-wide using the GENESIS software package [14], which applies generalized linear mixed models (GLMMs) for association testing while accounting for sample structure. GENESIS incorporates a kinship matrix to control for relatedness and population stratification, which we estimated separately within each ancestry group as described above. GWAS testing the association of SNP × HSV-1 interactions with AD risk and testing SNP associations with HSV-1 status conditional on AD status and covariates was conducted using fixed effect models including covarariates for age, sex, PCR status, tissue source, and varying number of global PCs that were significantly associated with each outcome model (Table S2). To improve interpretation of interactions, we also performed GWAS analyses in subgroups of individuals with and without detectable HSV-1. Analyses were conducted separately for each dataset and within ancestry group. Results across datasets and ancestry groups were combined using MR-MEGA [22], a meta-regression framework that models effect size heterogeneity across diverse ancestries. This approach extends inverse-variance weighted meta-analysis by incorporating study-specific principal components to account for ancestry-related heterogeneity.

We considered p-values < 0.05 significant for the effects of ε2 and ε4, and a standard genome-wide significance (GWS) threshold of p<5.0×10^−8^ for GWAS analyses. We also evaluated suggestive associations using a threshold of p < 5.0 × 10□□ to identify potentially relevant loci for follow-up. For the APOExHSV-1 interaction models, statistical inference was based on two-sided Wald tests derived from the estimated regression coefficients and their covariance matrix, with odds ratios and 95% confidence intervals reported on the exponentiated scale. Manhattan plots and QQ-Plots were generated using FUMA[23]. The web-based tools LDpair and LDexpress [24] were used to identify genes with expression levels associated with significant variants identified in the GWAS.

## 3. RESULTS

### 3.1 HSV-1 DNA is less prevalent in AD cases

DNA mapping to viral species sequences revealed that HSV-1 was the most highly prevalent virus, observed in 62.6% of EA, 80.1% of AA, 69.8% of CH, 65.8% of IND, 64.0% of EA/MID, and 87.8% of NAH individuals (Table S1). DNA from across the HSV-1 genome was detected in both blood and brain (Figure 1) and the majority mapped to the terminal and internal repeat long regions (IRL and TRL, respectively), which contain duplicated copies of the LAT locus, structurally and epigenetically dynamic domains implicated in recombination and latency related suppression of viral reactivation [100,101]. Table 1 shows the results of the association of detectable HSV-1 DNA with AD across ancestry groups and thresholds defining HSV-1 positivity. Significant associations showing a negative association between HSV-1 DNA and AD risk were observed in EA (OR=0.84, P=0.002), AA (OR=0.79, P=0.004), NAH (OR=0.62, P=1.18 × 10^−5^), and EA/MID (OR=0.63, P=0.001) ancestry groups, and a similar but not significant trend was observed in the CH (OR=0.86, P=0.06) and AJ (OR=0.81, P=0.17) groups. Effect directions were consistent in analyses stratified by blood/brain samples (Table S4).

**Figure 1.**
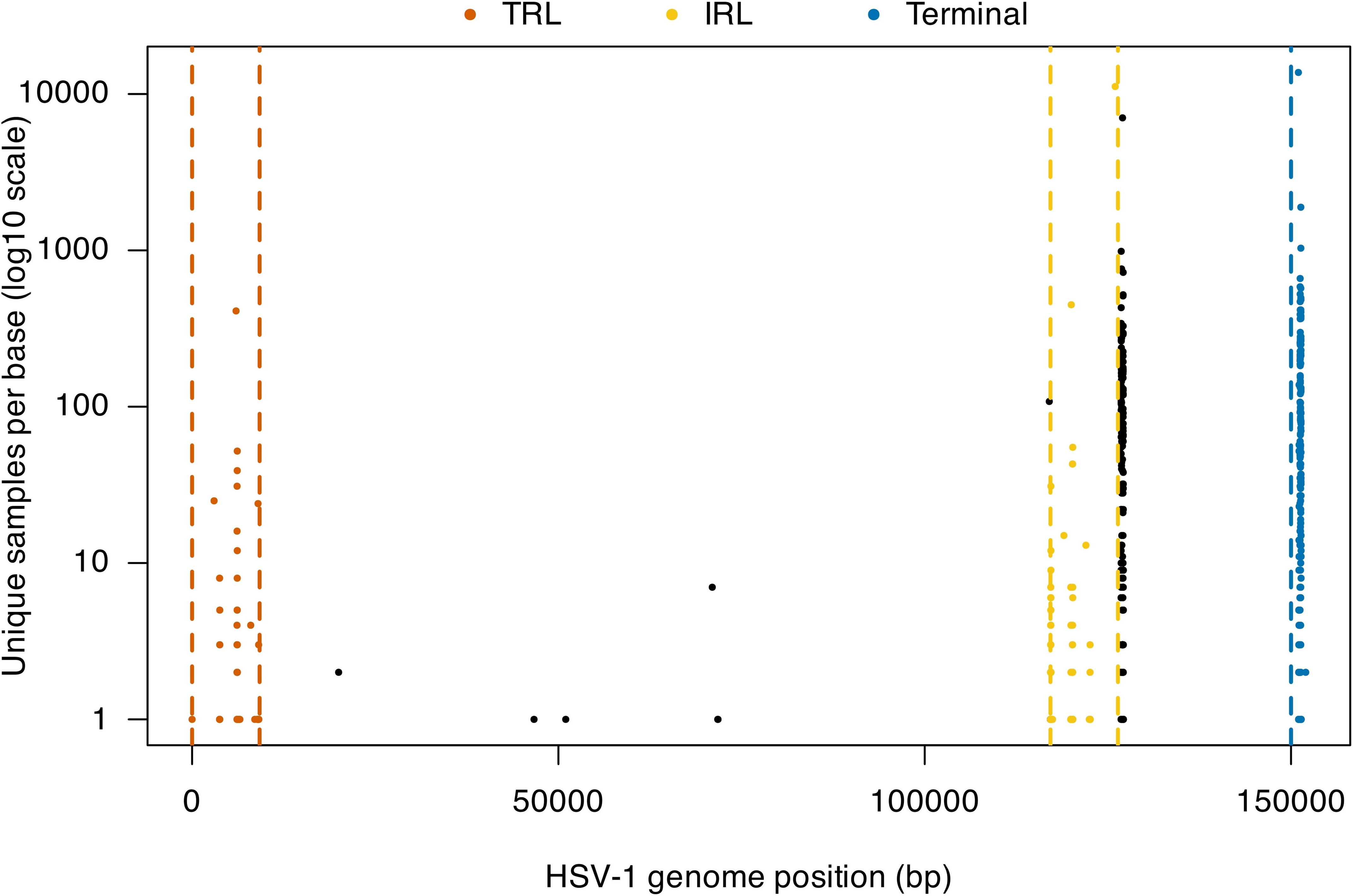
HSV-1 per-position sample density across the reference genome. Each point represents the number of unique samples with ≥1 HSV-1–aligned read at a genomic position (log10 scale). Points are colored by genomic region: TRL (terminal repeat long), IRL (internal repeat long; latency-associated transcript [LAT] locus, ∼17–126 kb), and the right terminal region (∼150 kb). Vertical dashed lines indicate boundaries of repeat long regions and the start of the right terminal region. Positions outside annotated regions are shown in black. Signal is enriched within repeat long regions containing the LAT locus and sparse across the unique long region.

### 3.2 Presence of HSV-1 DNA and its association with AD risk is *APOE* genotype-dependent

In the EA group, the odds of having detectable HSV-1 DNA was lower for *APOE-*ε4 carriers (OR = 0.83, P = 1.89×10^−4^) but higher for ε2 carriers (OR = 1.37, P = 4.96 × 10^−4^) compared to persons without those alleles (Table S5). The association among ε*2* carriers was evident in both controls (OR = 1.37, P = 0.007) and AD cases (OR = 1.41, P = 0.018). Among ε*4* carriers, the association was significant in controls (OR = 0.70, P = 2.81×10□□) and showed a similar but nonsignificant trend in AD cases (OR = 0.90, P = 0.11). Ancestry-specific patterns differed outside the EA group. In AA individuals, ε4 was linked to a higher odds of HSV-1 DNA detectability among AA controls (OR = 1.30, P = 0.02) but lower dectability of HSV-1 in EA/MID controls (OR = 0.49, P = 0.004) (Table S5). No significant *APOE* genotype effects were observed in other ancestry groups.

The association of AD with the interaction of ε4 and detectability HSV-1 DNA was significant and had similar magnitudes of effect in the EA (OR= 1.52, P =8.25 × 10^−5^) and CH (OR= 1.44, P = 0.02) groups. Further scrutiny of this relationship revealed that HSV-1 increased AD risk among ε4 carriers but was protective among persons lacking ε4 in both ancestry groups, noting that the protective effect among ε4 non-carriers was virtually identical in EA (OR = 0.72, P =6.58 × 10^−6^) and CH (OR = 0.80, P =0.04) individuals (Table 2). To isolate the contribution of HSV-1 beyond *APOE*-ε4, we compared the odds ratios for AD among ε4 carriers with HSV-1 (HSV1+/E4+) to those without HSV-1 (HSV1−/E4+). Detectable HSV-1 increased AD risk by 6.5% (2.38/2.17) in EA and 14% (1.94/1.70) in CH individuals in addition to the risk conferred by ε4 alone (Table 2). We performed a sensitivity analysis excluding ε2 carriers to determine whether the observed protective effect of detectable HSV-1 among ε4 non-carriers was attributable specifically to ε2. The ORs remained virtually unchanged in both EA (0.71) and CH (0.81) groups, suggesting that the observed protective effect of HSV-1 among ε4 non-carriers is independent of ε2 (Table S6).No significant virus × *APOE* interactions in the AA group or virus × ε2 interactions in any ancestry group were observed.

### 3.3 GWAS for detectable HSV-1 in human WGS data

There was little evidence of genomic inflation in the AA (λ = 1.006), AJ (λ = 1.006), EA/MID (λ = 1.016), EU (λ = 1.000), CH (λ = 1.007), and NAH (λ = 1.000) groups (Figures S2-5). In the CH group, we identified a GWS association of HSV-1 presence with *FAM227A* intronic SNP rs145502813 (OR = 1.60, P = 7.07 × 10□^10^) and suggestive associations with SNPs located between *TENT5* and *GDAP2, MTUS1*, and between *TCF3* and *ONECUT3* (Table 3, Figure S6). In the AA group, 11 correlated SNPs (r^2^ = 0.59, D’ = 0.83) located between *LINC02006* and *ARHGEF26* (ORs = 0.26-0.31, 4.78× 10□^8^ ≤ P ≤ 2.98 × 10□^8^) were significantly associated with lower odds of detectable HSV-1 DNA. Suggestive evidence for a protective effect against detectable HSV-1 was also observed with SNPs in *ZNF124*, between *SLC4A7* and *AC137675.1*, between *RARB* and *AC133141.1*, in *RARB*, and in *RBFOX1* (Table 3). Suggestive associations for increased odds of detecting HSV-1 DNA were also observed with SNPs in *TP63* in the AJ group and *LRFN5* in the EA group. No genome-wide significant or suggestive associations were detected in EA/MID, NAH, or IND groups.

No GWS associations were observed in the combined ancestry sample, although several suggestive associations were identified in *ADIRF-AS1,* near *AL157931.1* (P = 2.1–3.3 × 10□□) and in *CACNA2D1* (Table 4). In the combined sample of Caucasian population groups (EA, AJ, EA/MID), several GWS variants were observed including *CACNA2D1* rs10229296 P = 1.83×10^−8^) and rs10232780 (P = 2.08×10^−8^) with modest heterogeneity (P_het_ ≈ 0.04) and rs6014918 near *RP11-560A15.4* (P = 2.49 × 10□□) which is close to *BMP7*. Suggestive associations in this collective group were also detected with SNPs between *LPL* and *SLC18A1*, between *RP11-96C23.12* and *GLUD1*, and in *AP003108.2*, *LINC00353*, readthrough transcript *SYNJ2BP-COX16*, and *RBFOX1*.

### 3.4 GWAS for SNP x HSV-1 interactions affecting AD risk

Genome-wide scans for the association of AD with interactions between HSV-1 and SNPs revealed modest or moderate genomic inflation in the AA (λ = 1.006), AJ (λ = 0.99), EA/MID (λ = 1.12), EA (λ = 1.03), CH (λ = 1.07), and NAH (λ = 1.13) groups (Figures S7-S12). In the CH cohort, there was a GWS interaction with *C16orf74* SNP rs73257234 (OR = 3.30, P = 1.72 × 10□□) and a suggestive interaction with rs16880152 near *RNU6-663P* (Table 5, Figure S13). GWS interactions were observed with rs202096418 located between *CAPN11* and *SLC29A1* (OR = 0.24, P = 3.38 × 10□□) in the AJ group (Table 5, Figure S14) and *HOXD-AS2* SNP rs6718273 (OR = 0.33, P = 4.29 × 10□□) in the EA/MID group (Table 5, Figure S15). Suggestive signals in the EA/MID group included SNPs in *AC024559.2*, *EPHX2*, *ADAMTSL1*, and *CELF2*, as well as between *IL12RB2* and *SERBP1*. Suggestive evidence of interaction was observed with SNPs in *LOC105372064* and between *SH2D4B* and *LINC02655* in the AA cohort, with multiple *LINC00596* SNPs in the EA cohort, and between *AC068138.1 and COL4A3* in the NAH cohort (Table 5).

In the combined group of ancestry populations, a GWS interaction was observed with rs11690846 in *RBM45* (P = 4.35 × 10□□). Suggestive associations were detected with SNPs near *CDH2,* near *RP11-395B7.7,* between *RP11-388E23.3* and *LINC00596,* and in *MIR646HG*, *SCN1A*, *LINC01095*, and *TH2LCRR*. Results combined across the white populations revealed a GWS interaction with rs78893636 in *MGAT4C* (P = 3.54 × 10□□) and suggestive associations with SNPs between *CAPN11* and *SLC29A1*, between *TRIM56* and *SERPINE1*, and in *DPP6* and *KIAA1958* (Table 6).

Further analysis of GWS interactions revealed evidence of opposite effect directions by HSV-1 status (Table 5). In the AJ group, rs202096418 showed a protective effect in HSV-1 positive individuals (OR = 0.57, P = 0.001) but increased risk in HSV-1 negative individuals (OR = 2.54, P = 3.50 × 10□□). This same pattern was observed for rs6718273 in the EA/MID group (OR = 0.63, P = 2.27 × 10□□ in HSV-1 positive; OR = 1.81, P = 0.001 in HSV-1 negative). In the CH group, the rs73257234 effect allele was associated with increased AD risk among HSV-1 positive individuals (OR = 1.76, P = 6.27 × 10□□) but was protective in HSV-1 negative individuals (OR = 0.54, P = 4.08 × 10□□). None of the individual GWS association signals were significant in the combined ancestry populations.

## 4. DISCUSSION

### 4.1 Most HSV-1 DNA in ADSP participant tissue associated with inactive virus

In this study, we identified and quantified segments of HSV-1 DNA in whole genome sequence data from 27,315 AD cases and controls. The viral DNA sequences from nearly all individuals mapped to the HSV-1 terminal and internal repeat long regions (IRL and TRL). These regions contain duplicated copies of the latency-associated transcript and represent structurally and epigenetically dynamic domains implicated in recombination and latency related regulation of HSV-1 [25,26]. In contrast to conventional clinical assays for HSV-1, which detect active viral replication or serological responses, our sequencing-based approach identifies viral DNA independent of transcriptional or replicative activity. Our findings likely reflect differences in viral state and host control rather than timing of primary infection, as majority of detected HSV-1 DNA mapped to repeat regions containing the latency-associated transcript. Detection of HSV-1 DNA in blood therefore most likely reflects viral genomic material rather than productive replication and may represent latent episomes or transient reactivation. Most adults harbor lifelong HSV-1 latency in neurons, where chromatinized viral genomes remain transcriptionally silent and undetectable by sequencing [27], and individuals without detectable HSV-1 DNA may still carry deeply latent infection sequestered from immune surveillance [28]. In contrast, detectable viral DNA may capture more accessible or intermittently reactivating latent genomes [29,30].

### 4.2 Negative association between HSV-1 DNA and AD

Our finding that HSV-1 DNA is associated with lower risk of AD is novel and contradicts previous findings, including our earlier work using HSV-1 DNA [10], observational studies involving pathology in humans [31], and experimental evidence in animal models [1]. It should be noted that our previous finding of a positive association was observed in a study of primarily whole exome sequence data derived from blood samples from EA participants. Differences between whole exome and whole genome sequencing may influence the composition of detectable HSV-1 DNA. Capture-based and untargeted approaches differ between WES and WGS in coverage uniformity and off-target sequence recovery [32].

Although the effect direction we observed in the current study may appear counterintuitive, it was robust across blood and brain samples collected from individuals in multiple ancestry groups and sequenced in several laboratories. Rather than indicating a direct protective effect of HSV-1 infection, this pattern may reflect differences in viral latency and host control, because the majority of HSV-1 reads map to latency transcript regions. During latency, transcription from the LAT locus has been linked to repression of immediate early genes such as *ICP0*, a key transactivator required for lytic replication [33]. Little signal was observed across the unique long region, which encodes most genes required for active viral replication [34]. This pattern is consistent with detection of HSV-1 genomes in latent, rather than widespread lytic replication.There is some previous, albeit indirect evidence supporting a protective effect of HSV-1 in humans, however, including a recently completed clinical trial of the anti-viral agent Valacyclovir in mild AD patients that showed significantly poorer cognitive performance in the treatment group [31]. Another longitudinal PET imaging study found that HSV-1–positive individuals had lower cortical amyloid burden [35]. It is possible that HSV-1 infection, at least as measured by the presence of its DNA in a single sample of blood or brain tissue collected late in life of post-mortem, is associated with protection from AD pathology due to a beneficial immune response due to the presence of latent infections, although this is speculative and other explanations are possible.

### 4.2 HSV-1 detection in human DNA is genetically influenced and modifies genetic associatons with AD

This is the first study to show that the detectability of latent HSV-1 infection in human DNA is influenced by host genetics. Among EA individuals, *APOE* ε4 was associated with reduced odds of detectable HSV-1 DNA, whereas ε2 was associated with increased odds, patterns observed in both AD cases and controls. In contrast, *APOE* ε4 was associated with increased HSV-1 detectability among AA controls. Although detectable HSV-1 DNA was generally protective against AD, ε4 carriers showed the opposite pattern reduced detectability yet increased AD risk when HSV-1 was present. These findings suggest that ε4 may increase susceptibility to viral reactivation within the nervous system, promoting neurodegeneration even when peripheral detectability is low.

Our results align with prior reports that HSV-1 contributes to AD pathology primarily in ε4 carriers[36–37]. Mechanistic links are biologically plausible: HSV-1 glycoproteins bind heparan sulfate proteoglycans (HSPGs)[38], a high-affinity interaction site for *ApoE[*21]. *ApoE* ε4 binds more strongly to HSPGs than other isoforms[39–41], potentially enhancing viral neuronal entry and retention. Once reactivated in neurons, HSV-1 can trigger antimicrobial Aβ production[42], but impaired Aβ clearance in ε4 carriers may exacerbate harmful accumulation[43]. This idea is consistent with a study showing increased latent HSV-1 persistence in *APOE* ε4 mice, with ∼13-fold higher viral loads compared to *APOE* ε3 mice and markedly reduced latency in ApoE-deficient mice [44].

The protective association in ε4 non-carriers remains less well understood. Although ε2 can impair HSV-1 replication and attachment[21], our findings suggest additional immune or host-genetic influences beyond *APOE* alone. Stronger HSV-1 detectability in AA and CH individuals may reflect ancestry-specific differences in reactivation frequency or environmental exposures [45]. These observations underscore the importance of considering both ancestry and *APOE* background when evaluating the role of latent viral infections in AD.

### 4.3 Other SNPs Associated with Detectable HSV-1

Several SNPs associated with detectable viral DNA lie in or near genes relevant to immune regulation and neurodegeneration. *CACNA2D1* encodes a calcium channel subunit important for neuronal excitability[46], linking AD-related calcium dysregulation[47] with HSV-1 reliance on calcium entry pathways[48]. *BMP7* contributes in neuropeptide signaling changes induced by HSV-1 latency[49]. *GDAP2* contributes to cellular stress response and synaptic function[50]. *TCF3* is a regulator of Wnt/β-catenin signaling that influences neuronal development and cortical integrity[51,52]. Wnt signaling is critical for neuroprotection and blood–brain barrier function and is required for HSV-1 reactivation and active infection[51,53]. Genes involved in synaptic function further underscore convergence of viral and neurodegenerative pathways, including *RBFOX1* and *LRFN5* which affect neuronal activity and immune response[54–57], and reduced *RBFOX1* expression has been linked to amyloid burden and cognitive decline[57,58]. *TP63* regulates apoptosis, a process altered during HSV-1 infection[59].

### 4.4 Novel Interactions Between SNPs and Viral Presence Predict AD Risk

Genome-wide significant interactions include loci with biological links to AD. *MGAT4C* encodes an N-acetylglucosaminyltransferase involved in N-glycan branching, has been reported to be downregulated in AD brain [60], and variants in related glycosyltransferase genes *MGAT4B* and *MGAT5* have also been implicated as AD risk loci in GWAS meta-analyses [60–62]. *SLC29A1* (ENT1), which regulates adenosine signaling and blood–brain barrier integrity, influences HSV-1 replication—its inhibition enhances viral propagation [63,64]. Suggestive SNP × HSV-1 interactions were observed in genes related to immune and neuronal function, including GALNT1 involved in viral glycoprotein glycosylation [65], Although GALNT1 variants have not been directly linked to AD, GALNT1 protein levels correlate with visuospatial decline[66]. CDH2 supports synaptic structure [67], and the two genes at the *TRIM56/SERPINE1* locus have roles in antiviral and vascular inflammatory responses [68–71], and IL12RB2 which mediates Th1 signaling during HSV-1 infection[72–74], which has been associated with cognitive aging and AD risk[75].

### 4.5 Study limitations

This study leveraged a large, multi-ancestry WGS dataset with both blood and brain samples, but several limitations should be considered. Viral DNA detection reflects a single time point, limiting inference about temporality in relation to AD onset and does not allow inference about latent vs. active infections. Sequencing platforms and tissue sources varied across sites, although consistent HSV-1 detection across laboratories reduces concern for contamination. Power was reduced in some non-EA groups, and ancestry-specific heterogeneity may contribute to incomplete replication of findings. Finally, interpretation of SNP×HSV-1 interactions is constrained by smaller sample sizes within stratified analyses, and results require validation in independent cohorts.

## 5. Conclusion

Our results suggest that HSV-1 DNA detectability is negatively associated with AD risk in most individuals but confers increased risk in *APOE* ε4 carriers, and that host genetic variation influences both viral detectability and HSV-1–dependent genetic effects on AD. These findings highlight host–virus interactions as a potential contributor to heterogeneity in AD risk and warrant further investigation and replication. They also suggest that HSV-1 may have very different effects in the brain during latency compared to active/reactive infections.

## Supporting information

Main Tables

## Data Availability

All data can be obtained online at dbGaP.

https://dbgap.ncbi.nlm.nih.gov/home/

## ACKNOWLEDGEMENTS

*The Alzheimer’s Disease Sequencing Project (ADSP) is comprised of two Alzheimer’s Disease (AD) genetics consortia and three National Human Genome Research Institute (NHGRI) funded Large Scale Sequencing and Analysis Centers (LSAC). The two AD genetics consortia are the Alzheimer’s Disease Genetics Consortium (ADGC) funded by NIA (U01 AG032984), and the Cohorts for Heart and Aging Research in Genomic Epidemiology (CHARGE) funded by NIA (R01 AG033193), the National Heart, Lung, and Blood Institute (NHLBI), other National Institute of Health (NIH) institutes and other foreign governmental and non-governmental organizations. The Discovery Phase analysis of sequence data is supported through UF1AG047133 (to Drs. Schellenberg, Farrer, Pericak-Vance, Mayeux, and Haines); U01AG049505 to Dr. Seshadri; U01AG049506 to Dr. Boerwinkle; U01AG049507 to Dr. Wijsman; and U01AG049508 to Dr. Goate and the Discovery Extension Phase analysis is supported through U01AG052411 to Dr. Goate, U01AG052410 to Dr. Pericak-Vance and U01 AG052409 to Drs. Seshadri and Fornage.*

*Sequencing for the Follow Up Study (FUS) is supported through U01AG057659 (to Drs. PericakVance, Mayeux, and Vardarajan) and U01AG062943 (to Drs. Pericak-Vance and Mayeux). Data generation and harmonization in the Follow-up Phase is supported by U54AG052427 (to Drs. Schellenberg and Wang). The FUS Phase analysis of sequence data is supported through U01AG058589 (to Drs. Destefano, Boerwinkle, De Jager, Fornage, Seshadri, and Wijsman), U01AG058654 (to Drs. Haines, Bush, Farrer, Martin, and Pericak-Vance), U01AG058635 (to Dr. Goate), RF1AG058066 (to Drs. Haines, Pericak-Vance, and Scott), RF1AG057519 (to Drs. Farrer and Jun), R01AG048927 (to Dr. Farrer), and RF1AG054074 (to Drs. Pericak-Vance and Beecham).*

*The ADGC cohorts include: Adult Changes in Thought (ACT) (U01 AG006781, U19 AG066567), the Alzheimer’s Disease Research Centers (ADRC) (P30 AG062429, P30 AG066468, P30 AG062421, P30 AG066509, P30 AG066514, P30 AG066530, P30 AG066507, P30 AG066444, P30 AG066518, P30 AG066512, P30 AG066462, P30 AG072979, P30 AG072972, P30 AG072976, P30 AG072975, P30 AG072978, P30 AG072977, P30 AG066519, P30 AG062677, P30 AG079280, P30 AG062422, P30 AG066511, P30 AG072946, P30 AG062715, P30 AG072973, P30 AG066506, P30 AG066508, P30 AG066515, P30 AG072947, P30 AG072931, P30 AG066546, P20 AG068024, P20 AG068053, P20 AG068077, P20 AG068082, P30 AG072958, P30 AG072959), the Chicago Health and Aging Project (CHAP) (R01 AG11101, RC4 AG039085, K23 AG030944), Indiana Memory and Aging Study (IMAS) (R01 AG019771), Indianapolis Ibadan (R01 AG009956, P30 AG010133), the Memory and Aging Project (MAP) (R01 AG17917), Mayo Clinic (MAYO) (R01 AG032990, U01 AG046139, R01 NS080820, RF1 AG051504, P50 AG016574), Mayo Parkinson’s Disease controls (NS039764, NS071674, 5RC2HG005605), University of Miami (R01 AG027944, R01 AG028786, R01 AG019085, IIRG09133827, A2011048), the Multi-Institutional Research in Alzheimer’s Genetic Epidemiology Study (MIRAGE) (R01 AG09029, R01 AG025259), the National Centralized Repository for Alzheimer’s Disease and Related Dementias (NCRAD) (U24 AG021886), the National Institute on Aging Late Onset Alzheimer’s Disease Family Study (NIA- LOAD) (U24 AG056270), the Religious Orders Study (ROS) (P30 AG10161, R01 AG15819), the Texas Alzheimer’s Research and Care Consortium (TARCC) (funded by the Darrell K Royal Texas Alzheimer’s Initiative), Vanderbilt University/Case Western Reserve University (VAN/CWRU) (R01 AG019757, R01 AG021547, R01 AG027944, R01 AG028786, P01 NS026630, and Alzheimer’s Association), the Washington Heights-Inwood Columbia Aging Project (WHICAP) (RF1 AG054023), the University of Washington Families (VA Research Merit Grant, NIA: P50AG005136, R01AG041797, NINDS: R01NS069719), the Columbia University Hispanic Estudio Familiar de Influencia Genetica de Alzheimer (EFIGA) (RF1 AG015473), the University of Toronto (UT) (funded by Wellcome Trust, Medical Research Council, Canadian Institutes of Health Research), and Genetic Differences (GD) (R01 AG007584). The CHARGE cohorts are supported in part by National Heart, Lung, and Blood Institute (NHLBI) infrastructure grant HL105756 (Psaty), RC2HL102419 (Boerwinkle) and the neurology working group is supported by the National Institute on Aging (NIA) R01 grant AG033193*.

*The CHARGE cohorts participating in the ADSP include the following: Austrian Stroke Prevention Study (ASPS), ASPS-Family study, and the Prospective Dementia Registry-Austria (ASPS/PRODEM-Aus), the Atherosclerosis Risk in Communities (ARIC) Study, the Cardiovascular Health Study (CHS), the Erasmus Rucphen Family Study (ERF), the Framingham Heart Study (FHS), and the Rotterdam Study (RS). ASPS is funded by the Austrian Science Fond (FWF) grant number P20545-P05 and P13180 and the Medical University of Graz. The ASPS-Fam is funded by the Austrian Science Fund (FWF) project I904), the EU Joint Programme – Neurodegenerative Disease Research (JPND) in frame of the BRIDGET project (Austria, Ministry of Science) and the Medical University of Graz and the Steiermärkische Krankenanstalten Gesellschaft. PRODEM-Austria is supported by the Austrian Research Promotion agency (FFG) (Project No. 827462) and by the Austrian National Bank (Anniversary Fund, project 15435. ARIC research is carried out as a collaborative study supported by NHLBI contracts (HHSN268201100005C, HHSN268201100006C, HHSN268201100007C, HHSN268201100008C, HHSN268201100009C, HHSN268201100010C, HHSN268201100011C, and HHSN268201100012C). Neurocognitive data in ARIC is collected by U01 2U01HL096812, 2U01HL096814, 2U01HL096899, 2U01HL096902, 2U01HL096917 from the NIH (NHLBI, NINDS, NIA and NIDCD), and with previous brain MRI examinations funded by R01-HL70825 from the NHLBI. CHS research was supported by contracts HHSN268201200036C, HHSN268200800007C, N01HC55222, N01HC85079, N01HC85080, N01HC85081, N01HC85082, N01HC85083, N01HC85086, and grants U01HL080295 and U01HL130114 from the NHLBI with additional contribution from the National Institute of Neurological Disorders and Stroke (NINDS). Additional support was provided by R01AG023629, R01AG15928, and R01AG20098 from the NIA. FHS research is supported by NHLBI contracts N01-HC-25195 and HHSN268201500001I. This study was also supported by additional grants from the NIA (R01s AG054076, AG049607 and AG033040 and NINDS (R01 NS017950). The ERF study as a part of EUROSPAN (European Special Populations Research Network) was supported by European Commission FP6 STRP grant number 018947 (LSHG-CT-2006-01947) and also received funding from the European Community’s Seventh Framework Programme (FP7/2007-2013)/grant agreement HEALTH-F4- 2007-201413 by the European Commission under the programme “Quality of Life and Management of the Living Resources” of 5th Framework Programme (no. QLG2-CT-2002- 01254). High-throughput analysis of the ERF data was supported by a joint grant from the Netherlands Organization for Scientific Research and the Russian Foundation for Basic Research (NWO-RFBR 047.017.043). The Rotterdam Study is funded by Erasmus Medical Center and Erasmus University, Rotterdam, the Netherlands Organization for Health Research and Development (ZonMw), the Research Institute for Diseases in the Elderly (RIDE), the Ministry of Education, Culture and Science, the Ministry for Health, Welfare and Sports, the European Commission (DG XII), and the municipality of Rotterdam. Genetic data sets are also supported by the Netherlands Organization of Scientific Research NWO Investments (175.010.2005.011, 911-03-012), the Genetic Laboratory of the Department of Internal Medicine, Erasmus MC, the Research Institute for Diseases in the Elderly (014-93-015; RIDE2), and the Netherlands Genomics Initiative (NGI)/Netherlands Organization for Scientific Research (NWO) Netherlands Consortium for Healthy Aging (NCHA), project 050-060-810. All studies are grateful to their participants, faculty and staff. The content of these manuscripts is solely the responsibility of the authors and does not necessarily represent the official views of the National Institutes of Health or the U.S. Department of Health and Human Services*.

*The FUS cohorts include: the Alzheimer’s Disease Research Centers (ADRC) (P30 AG062429, P30 AG066468, P30 AG062421, P30 AG066509, P30 AG066514, P30 AG066530, P30 AG066507, P30 AG066444, P30 AG066518, P30 AG066512, P30 AG066462, P30 AG072979, P30 AG072972, P30 AG072976, P30 AG072975, P30 AG072978, P30 AG072977, P30 AG066519, P30 AG062677, P30 AG079280, P30 AG062422, P30 AG066511, P30 AG072946, P30 AG062715, P30 AG072973, P30 AG066506, P30 AG066508, P30 AG066515, P30 AG072947, P30 AG072931, P30 AG066546, P20 AG068024, P20 AG068053, P20 AG068077, P20 AG068082, P30 AG072958, P30 AG072959), Alzheimer’s Disease Neuroimaging Initiative (ADNI) (U19AG024904), Amish Protective Variant Study (RF1AG058066), Cache County Study (R01AG11380, R01AG031272, R01AG21136, RF1AG054052), Case Western Reserve University Brain Bank (CWRUBB) (P50AG008012), Case Western Reserve University Rapid Decline (CWRURD) (RF1AG058267, NU38CK000480), CubanAmerican Alzheimer’s Disease Initiative (CuAADI) (3U01AG052410), Estudio Familiar de Influencia Genetica en Alzheimer (EFIGA) (5R37AG015473, RF1AG015473, R56AG051876), Genetic and Environmental Risk Factors for Alzheimer Disease Among African Americans Study (GenerAAtions) (2R01AG09029, R01AG025259, 2R01AG048927), Gwangju Alzheimer and Related Dementias Study (GARD) (U01AG062602), Hillblom Aging Network (2014-A-004-NET, R01AG032289, R01AG048234), Hussman Institute for Human Genomics Brain Bank (HIHGBB) (R01AG027944, Alzheimer’s Association “Identification of Rare Variants in Alzheimer Disease”), Ibadan Study of Aging (IBADAN) (5R01AG009956), Longevity Genes Project (LGP) and LonGenity (R01AG042188, R01AG044829, R01AG046949, R01AG057909, R01AG061155, P30AG038072), Mexican Health and Aging Study (MHAS) (R01AG018016), Multi-Institutional Research in Alzheimer’s Genetic Epidemiology (MIRAGE) (2R01AG09029, R01AG025259, 2R01AG048927), Northern Manhattan Study (NOMAS) (R01NS29993), Peru Alzheimer’s Disease Initiative (PeADI) (RF1AG054074), Puerto Rican 1066 (PR1066) (Wellcome Trust (GR066133/GR080002), European Research Council (340755)), Puerto Rican Alzheimer Disease Initiative (PRADI) (RF1AG054074), Reasons for Geographic and Racial Differences in Stroke (REGARDS) (U01NS041588), Research in African American Alzheimer Disease Initiative (REAAADI) (U01AG052410), the Religious Orders Study (ROS) (P30 AG10161, P30 AG72975, R01 AG15819, R01 AG42210), the RUSH Memory and Aging Project (MAP) (R01 AG017917, R01 AG42210Stanford Extreme Phenotypes in AD (R01AG060747), University of Miami Brain Endowment Bank (MBB), University of Miami/Case Western/North Carolina A&T African American (UM/CASE/NCAT) (U01AG052410, R01AG028786), Wisconsin Registry for Alzheimer’s Prevention (WRAP) (R01AG027161 and R01AG054047), Mexico-Southern California Autosomal Dominant Alzheimer’s Disease Consortium (R01AG069013), Center for Cognitive Neuroscience and Aging (R01AG047649), and the A4 Study (R01AG063689, U19AG010483 and U24AG057437)*.

*The four LSACs are: the Human Genome Sequencing Center at the Baylor College of Medicine (U54 HG003273), the Broad Institute Genome Center (U54HG003067), The American Genome Center at the Uniformed Services University of the Health Sciences (U01AG057659), and the Washington University Genome Institute (U54HG003079*). *Genotyping and sequencing for the ADSP FUS is also conducted at John P. Hussman Institute for Human Genomics (HIHG) Center for Genome Technology (CGT)*.

*Biological samples and associated phenotypic data used in primary data analyses were stored at Study Investigators institutions, and at the National Centralized Repository for Alzheimer’s Disease and Related Dementias (NCRAD, U24AG021886) at Indiana University funded by NIA. Associated Phenotypic Data used in primary and secondary data analyses were provided by Study Investigators, the NIA funded Alzheimer’s Disease Centers (ADCs), and the National Alzheimer’s Coordinating Center (NACC, U24AG072122) and the National Institute on Aging Genetics of Alzheimer’s Disease Data Storage Site (NIAGADS, U24AG041689) at the University of Pennsylvania, funded by NIA. Harmonized phenotypes were provided by the ADSP Phenotype Harmonization Consortium (ADSP-PHC), funded by NIA (U24 AG074855, U01 AG068057 and R01 AG059716) and Ultrascale Machine Learning to Empower Discovery in Alzheimer’s Disease Biobanks (AI4AD, U01 AG068057). This research was supported in part by the Intramural Research Program of the National Institutes of health, National Library of Medicine. Contributors to the Genetic Analysis Data included Study Investigators on projects that were individually funded by NIA, and other NIH institutes, and by private U.S. organizations, or foreign governmental or nongovernmental organizations*.

The ADSP Phenotype Harmonization Consortium (ADSP-PHC) is funded by NIA (U24 AG074855, U01 AG068057 and R01 AG059716). The harmonized cohorts within the ADSP-PHC include: the Anti-Amyloid Treatment in Asymptomatic Alzheimer’s study (A4 Study), a secondary prevention trial in preclinical Alzheimer’s disease, aiming to slow cognitive decline associated with brain amyloid accumulation in clinically normal older individuals. The A4 Study is funded by a public-private-philanthropic partnership, including funding from the National Institutes of Health-National Institute on Aging, Eli Lilly and Company, Alzheimer’s Association, Accelerating Medicines Partnership, GHR Foundation, an anonymous foundation and additional private donors, with in-kind support from Avid and Cogstate. The companion observational Longitudinal Evaluation of Amyloid Risk and Neurodegeneration (LEARN) Study is funded by the Alzheimer’s Association and GHR Foundation. The A4 and LEARN Studies are led by Dr. Reisa Sperling at Brigham and Women’s Hospital, Harvard Medical School and Dr. Paul Aisen at the Alzheimer’s Therapeutic Research Institute (ATRI), University of Southern California. The A4 and LEARN Studies are coordinated by ATRI at the University of Southern California, and the data are made available through the Laboratory for Neuro Imaging at the University of Southern California. The participants screening for the A4 Study provided permission to share their de-identified data in order to advance the quest to find a successful treatment for Alzheimer’s disease. We would like to acknowledge the dedication of all the participants, the site personnel, and all of the partnership team members who continue to make the A4 and LEARN Studies possible. The complete A4 Study Team list is available on: a4study.org/a4-study-team.; the Adult Changes in Thought study (ACT), *U01 AG006781, U19 AG066567;* Alzheimer’s Disease Neuroimaging Initiative (ADNI): *Data collection and sharing for this project was funded by the Alzheimer’s Disease Neuroimaging Initiative (ADNI) (National Institutes of Health Grant U01 AG024904) and DOD ADNI (Department of Defense award number W81XWH-12-2-0012). ADNI is funded by the National Institute on Aging, the National Institute of Biomedical Imaging and Bioengineering, and through generous contributions from the following: AbbVie, Alzheimer’s Association; Alzheimer’s Drug Discovery Foundation; Araclon Biotech; BioClinica, Inc.; Biogen; Bristol-Myers Squibb Company; CereSpir, Inc.; Cogstate; Eisai Inc.; Elan Pharmaceuticals, Inc.; Eli Lilly and Company; EuroImmun; F. Hoffmann-La Roche Ltd and its affiliated company Genentech, Inc.; Fujirebio; GE Healthcare; IXICO Ltd.;Janssen Alzheimer Immunotherapy Research & Development, LLC.; Johnson & Johnson Pharmaceutical Research & Development LLC.; Lumosity; Lundbeck; Merck & Co., Inc.;Meso Scale Diagnostics, LLC.; NeuroRx Research; Neurotrack Technologies; Novartis Pharmaceuticals Corporation; Pfizer Inc.; Piramal Imaging; Servier; Takeda Pharmaceutical Company; and Transition Therapeutics. The Canadian Institutes of Health Research is providing funds to support ADNI clinical sites in Canada. Private sector contributions are facilitated by the Foundation for the National Institutes of Health (*www.fnih.org*). The grantee organization is the Northern California Institute for Research and Education, and the study is coordinated by the Alzheimer’s Therapeutic Research Institute at the University of Southern California. ADNI data are disseminated by the Laboratory for Neuro Imaging at the University of Southern California;* Estudio Familiar de Influencia Genetica en Alzheimer (EFIGA): *5R37AG015473, RF1AG015473, R56AG051876;* the Health & Aging Brain Study – Health Disparities (HABS-HD), supported by the National Institute on Aging of the National Institutes of Health under Award Numbers R01AG054073, R01AG058533, R01AG070862, P41EB015922, and U19AG078109; the Korean Brain Aging Study for the Early Diagnosis and Prediction of Alzheimer’s disease (KBASE), which was supported by a grant from Ministry of Science, ICT and Future Planning (Grant No: NRF-2014M3C7A1046042); Memory & Aging Project at Knight Alzheimer’s Disease Research Center (MAP at Knight ADRC): The Memory and Aging Project at the Knight-ADRC (Knight-ADRC). This work was supported by the National Institutes of Health (NIH) grants R01AG064614, R01AG044546, RF1AG053303, RF1AG058501, U01AG058922 and R01AG064877 to Carlos Cruchaga. The recruitment and clinical characterization of research participants at Washington University was supported by NIH grants P30AG066444, P01AG03991, and P01AG026276. Data collection and sharing for this project was supported by NIH grants RF1AG054080, P30AG066462, R01AG064614 and U01AG052410. We thank the contributors who collected samples used in this study, as well as patients and their families, whose help and participation made this work possible. This work was supported by access to equipment made possible by the Hope Center for Neurological Disorders, the Neurogenomics and Informatics Center (NGI: https://neurogenomics.wustl.edu/) and the Departments of Neurology and Psychiatry at Washington University School of Medicine; National Alzheimer’s Coordinating Center (NACC): *The NACC database is funded by NIA/NIH Grant U24 AG072122.* SCAN is a multi-institutional project that was funded as a U24 grant (AG067418) by the National Institute on Aging in May 2020. Data collected by SCAN and shared by NACC are contributed by the NIA-funded ADRCs as follows*: P30 AG062429 (PI James Brewer, MD, PhD), P30 AG066468 (PI Oscar Lopez, MD), P30 AG062421 (PI Bradley Hyman, MD, PhD), P30 AG066509 (PI Thomas Grabowski, MD), P30 AG066514 (PI Mary Sano, PhD), P30 AG066530 (PI Helena Chui, MD), P30 AG066507 (PI Marilyn Albert, PhD), P30 AG066444 (PI John Morris, MD), P30 AG066518 (PI Jeffrey Kaye, MD), P30 AG066512 (PI Thomas Wisniewski, MD), P30 AG066462 (PI Scott Small, MD), P30 AG072979 (PI David Wolk, MD), P30 AG072972 (PI Charles DeCarli, MD), P30 AG072976 (PI Andrew Saykin, PsyD), P30 AG072975 (PI David Bennett, MD), P30 AG072978 (PI Neil Kowall, MD), P30 AG072977 (PI Robert Vassar, PhD), P30 AG066519 (PI Frank LaFerla, PhD), P30 AG062677 (PI Ronald Petersen, MD, PhD), P30 AG079280 (PI Eric Reiman, MD), P30 AG062422 (PI Gil Rabinovici, MD), P30 AG066511 (PI Allan Levey, MD, PhD), P30 AG072946 (PI Linda Van Eldik, PhD), P30 AG062715 (PI Sanjay Asthana, MD, FRCP), P30 AG072973 (PI Russell Swerdlow, MD), P30 AG066506 (PI Todd Golde, MD, PhD), P30 AG066508 (PI Stephen Strittmatter, MD, PhD), P30 AG066515 (PI Victor Henderson, MD, MS), P30 AG072947 (PI Suzanne Craft, PhD), P30 AG072931 (PI Henry Paulson, MD, PhD), P30 AG066546 (PI Sudha Seshadri, MD), P20 AG068024 (PI Erik Roberson, MD, PhD), P20 AG068053 (PI Justin Miller, PhD), P20 AG068077 (PI Gary Rosenberg, MD), P20 AG068082 (PI Angela Jefferson, PhD), P30 AG072958 (PI Heather Whitson, MD), P30 AG072959 (PI James Leverenz, MD);* National Institute on Aging Alzheimer’s Disease Family Based Study (NIA-AD FBS): U24 AG056270; Religious Orders Study (ROS): P30AG10161,R01AG15819, R01AG42210; Memory and Aging Project (MAP - Rush): R01AG017917, R01AG42210; Minority Aging Research Study (MARS): R01AG22018, R01AG42210; the Texas Alzheimer’s Research and Care Consortium (TARCC), funded by the Darrell K Royal Texas Alzheimer’s Initiative, directed by the Texas Council on Alzheimer’s Disease and Related Disorders; Washington Heights/Inwood Columbia Aging Project (WHICAP): *RF1 AG054023;*and Wisconsin Registry for Alzheimer’s Prevention (WRAP): *R01AG027161 and R01AG054047.* Additional acknowledgments include the National Institute on Aging Genetics of Alzheimer’s Disease Data Storage Site (NIAGADS, U24AG041689) at the University of Pennsylvania, funded by NIA.

## CONFLICTS OF INTEREST

The authors declare that they have no competing interests.

## FUNDING

Data used in the preparation of this article were obtained from the Alzheimer’s Disease Sequencing Project (ADSP), funded by the National Institute on Aging (NIA) and the National Human Genome Research Institute (NHGRI). The funders had no role in study design, data analysis, interpretation of results, or manuscript preparation.

## CONSENT STATEMENT

All human data used in this study were obtained from controlled-access repositories. The original studies from which these data were derived were approved by their respective Institutional Review Boards, and all participants or their legally authorized representatives provided written informed consent for genetic research and data sharing. This study involved secondary analysis of de-identified data and was therefore exempt from additional institutional review board approval.

## Notes

### Competing Interest Statement

The authors have declared no competing interest.

### Funding Statement

Data used in the preparation of this article were obtained from the Alzheimers Disease Sequencing Project (ADSP), funded by the National Institute on Aging (NIA) and the National Human Genome Research Institute (NHGRI). The funders had no role in study design, data analysis, interpretation of results, or manuscript preparation.

