## Supplementary material for "Herpes simplex virus type 1 DNA is less prevalent in persons with Alzheimer’s disease and genetic factors modify the effect": Main Tables

**Table 1. Association between HSV-1 DNA and AD by ancestry across multiple read count thresholds**

| **Ancestry** | **HSV-1 > 0** | | **HSV-1 > 1** | | **HSV-1 >= 5** | | **HSV-1 > = 20** | |
| --- | --- | --- | --- | --- | --- | --- | --- | --- |
|  | **Odds Ratio** | **P-value** | **Odds Ratio** | **P-value** | **Odds Ratio** | **P-value** | **Odds Ratio** | **P-value** |
| AA | 0.79 | 0.004 | 0.79 | 0.005 | 0.85 | 0.020 | 0.81 | 0.052 |
| AJ | 0.81 | 0.166 | 0.81 | 0.166 | 0.88 | 0.415 | 0.72 | 0.285 |
| CH | 0.88 | 0.202 | 0.88 | 0.202 | 0.86 | 0.065 | 0.95 | 0.521 |
| EA | 0.84 | 0.002 | 0.84 | 0.002 | 0.70 | 1.45x10^-10^ | 0.90 | 0.272 |
| IND | 1.29 | 0.238 | 1.29 | 0.238 | 0.77 | 0.273 | 1.01 | 0.984 |
| NAH | 0.62 | 1.18x10^-5^ | 0.62 | 1.41x10^-5^ | 0.64 | 5.62x10^-8^ | 0.77 | 0.012 |
| EA/MID | 0.63 | 0.001 | 0.63 | 0.001 | 0.52 | 3.39x10^-6^ | 0.67 | 0.112 |

**Table 2.** Joint effects of HSV-1 and the *APOE* ɛ4 allele on AD risk by ancestry group

| **Ancestry^*^** | **HSV-1 and *APOE* ɛ4 Status** | **Odds Ratio** ^†^ | **CI^‡^** | **P-value** |
| --- | --- | --- | --- | --- |
| EA | HSV1(+)/E4(+) | 2.38 | [2.04, 2.77] | 1.94x10^-28^ |
|  | HSV1(+)/E4(-) | 0.72 | [0.63, 0.83] | 6.58x10^-6^ |
|  | HSV1(-)/E4(+) | 2.17 | [1.84, 2.55] | 1.38x10^-20^ |
|  | HSV1(-)/E4(-) | 1.00 | - | - |
| CH | HSV1(+)/E4(+) | 1.94 | [1.52, 2.47] | 1.06x10^-7^ |
|  | HSV1(+)/E4(-) | 0.80 | [0.64, 0.99] | 0.04 |
|  | HSV1(-)/E4(+) | 1.70 | [1.32, 2.17] | 2.95x10^-5^ |
|  | HSV1(-)/E4(-) | 1.00 | - | - |

^*^ EA = European ancestry; CH = Caribbean Hispanics

^†^ compared to the HSV1(-) / **ɛ**4(-) referent group

^‡^ CI = 95% confidence interval

**Table 3. Top-ranked associations (P < 5.0x10^-7^) for HSV-1 DNA presence by ancestry group**

| **Ancestry^*^** | **Chr** | **Position** | **SNP** | **Gene or Locus** | **MAF^†^** | **OR^‡^** | **P-value** |
| --- | --- | --- | --- | --- | --- | --- | --- |
| AA | 1 | 247154331 | rs61837909 | ZNF124 | 0.013 | 0.31 | 4.29x10^-7^ |
|  | 3 | 25483204 | rs76594842 | RARB | 0.040 | 0.50 | 3.91x10^-7^ |
|  |  | 27563891 | rs34345115 | SLC4A7 // AC137675.1 | 0.101 | 0.63 | 3.91x10^-7^ |
|  |  | 86442777 | rs73844213 | RARB// AC133141.1 | 0.107 | 0.64 | 3.18x10^-7^ |
|  |  | 153864584 | rs182529481 | LINC02006// ARHGEF26 | 0.013 | 0.26 | 2.98x10^-8^ |
|  |  | 153890997 | rs147751408 | AC068985.1 | 0.014 | 0.31 | 4.93x10^-7^ |
|  | 16 | 6727498 | rs114690150 | RBFOX1 | 0.081 | 0.61 | 4.76x10^-7^ |
| AJ | 3 | 189714643 | rs11915751 | TP63 | 0.687 | 1.66 | 4.17x10^-7^ |
| CH | 1 | 117748943 | rs11577247 | TENT5C/GDAP2 | 0.770 | 0.73 | 2.07x10^-7^ |
|  | 8 | 17718557 | rs74327930 | MTUS1 | 0.021 | 0.42 | 3.46x10^-7^ |
|  | 19 | 1711987 | rs112299149 | TCF3/ONECUT3 | 0.473 | 0.77 | 2.74x10^-7^ |
|  | 22 | 38644593 | rs145502813 | FAM227A | 0.143 | 1.60 | 7.07x10^-10^ |
| EA | 14 | 41878204 | rs72670419 | LRFN5 | 0.230 | 1.24 | 1.39x10^-7^ |

^*^ AA = African American, AJ = Ashkenazi Jewish, CH = Caribbean Hispanic, EA = European ancestry

^†^ MAF = minor allele frequency

^‡^ OR = Odds ratio

**Table 4. Top-ranked associations (P < 5.0x10^-7^) for HSV-1 DNA presence by meta-analysis group**

| **Study Group** | **Chr** | **Locus** | **SNP** | **Gene** | **P-meta** | **P-het** | **P-resid** | **Dir** |
| --- | --- | --- | --- | --- | --- | --- | --- | --- |
| ALL | 7 | 81851811 | rs10229296 | CACNA2D1 | 9.14x10^-8^ | 6.00x10^-7^ | 0.063 | +++--+ |
|  |  | 81852118 | rs10232780 | CACNA2D1 | 1.74x10^-7^ | 2.10x10^-6^ | 0.064 | +++--+ |
|  | 10 | 88727707 | rs1240382 | ADIRF-AS1 | 3.60x10^-7^ | 3.74x10^-6^ | 0.426 | ---+-+ |
|  |  | 89137271 | rs2782077 | NPAP1P3 | 3.54x10^-7^ | 3.68x10^-7^ | 0.925 | +++--- |
|  | 13 | 23560650 | rs9510486 | AL157931.1 | 2.07x10^-7^ | 0.001 | 0.722 | -----+- |
|  |  | 23563089 | rs7983789 | AL157931.1 | 3.29x10^-7^ | 0.001 | 0.703 | -----+- |
|  |  | 23563893 | rs9580495 | AL157931.1 | 2.94x10^-7^ | 0.001 | 0.721 | -----+- |
| EUR | 7 | 81851811 | rs10229296 | CACNA2D1 | 1.83x10^-8^ | 0.044 | 0.167 | +++ |
|  |  | 81852118 | rs10232780 | CACNA2D1 | 2.08x10^-8^ | 0.043 | 0.155 | +++ |
|  | 8 | 19854773 | rs2410622 | LPL//SLC18A1 | 1.85x10^-7^ | 0.044 | 0.852 | --- |
|  | 10 | 88727079 | rs1240387 | RP11-96C23.12/ GLUD1 | 4.48x10^-7^ | 0.097 | 0.516 | --- |
|  | 11 | 61167109 | rs4939521 | AP003108.2 | 4.18x10^-7^ | 0.009 | 0.555 | +++ |
|  | 13 | 90203131 | rs80001466 | LINC00353 | 2.90x10^-7^ | 0.002 | 0.509 | +++ |
|  | 14 | 70811990 | rs61977514 | SYNJ2BP-COX16 | 4.40x10^-7^ | 0.865 | 0.878 | --- |
|  | 16 | 6727498 | rs116986667 | RBFOX1 | 4.42x10^-7^ | 0.001 | 0.235 | --- |
|  | 20 | 55662514 | rs6014918 | RP11-560A15.4 / near BMP7 | 2.49x10^-8^ | 0.264 | 0.715 | +++ |

**Table 5. Genome-wide significant and suggestive (P < 5.0x10^-7^) associations of SNP × HSV-1 interaction with AD risk**

| **Ancestry^*^** | **Chr** | **Position** | **SNP** | **Gene** | **Model^†^** | **MAF^‡^** | **OR^§^** | **P-value** |
| --- | --- | --- | --- | --- | --- | --- | --- | --- |
| AA | 10 | 80674270 | rs143251710 | SH2D4B//LINC02655 | Interaction | 0.02 | 0.09 | 4.07x10^-7^ |
|  |  |  |  |  | HSV-1+ | 0.021 | 0.48 | 5.28x10^-5^ |
|  |  |  |  |  | HSV-1 - | 0.013 | 5.11 | 4.97x10^-4^ |
|  | 18 | 35730152 | rs116135624 | LOC105372064 | Interaction | 0.042 | 4.24 | 2.55x10^-7^ |
|  |  |  |  |  | HSV-1+ | 0.042 | 1.51 | 0.002 |
|  |  |  |  |  | HSV-1 - | 0.041 | 0.35 | 8.67x10^-5^ |
| AJ | 6 | 44195472 | rs202096418 | CAPN11 // SLC29A1 | Interaction | 0.205 | 0.24 | 3.38x10^-8^ |
|  |  |  |  |  | HSV-1+ | 0.198 | 0.57 | 0.001 |
|  |  |  |  |  | HSV-1 - | 0.215 | 2.54 | 3.50x10^-6^ |
| CH | 8 | 32904194 | rs16880152 | RNU6-663P | Interaction | 0.014 | 7.13 | 2.93x10^-7^ |
|  |  |  |  |  | HSV-1+ | 0.011 | 3.77 | 1.50x10^-7^ |
|  |  |  |  |  | HSV-1 - | 0.018 | 0.55 | 0.048 |
|  | 16 | 85742990 | rs73257234 | C16orf74 | Interaction | 0.045 | 3.3 | 1.72x10^-8^ |
|  |  |  |  |  | HSV-1+ | 0.038 | 1.76 | 6.27x10^-5^ |
|  |  |  |  |  | HSV-1 - | 0.059 | 0.54 | 4.08x10^-4^ |
| EA/MID | 1 | 67469123 | rs72934933 | IL12RB2//SERBP1 | Interaction | 0.03 | 17.95 | 4.26x10^-7^ |
|  |  |  |  |  | HSV-1+ | 0.031 | 2.53 | 0.006 |
|  |  |  |  |  | HSV-1 - | 0.027 | 0.15 | 2.01x10^-4^ |
|  | 2 | 176127142 | rs6718273 | HOXD-AS2 | Interaction | 0.632 | 0.33 | 4.29x10^-8^ |
|  |  |  |  |  | HSV-1+ | 0.631 | 0.63 | 2.27x10^-4^ |
|  |  |  |  |  | HSV-1 - | 0.632 | 1.81 | 0.001 |
|  | 3 | 194086501 | rs13099626 | AC024559.2 | Interaction | 0.399 | 0.37 | 2.02x10^-7^ |
|  |  |  |  |  | HSV-1+ | 0.4 | 0.65 | 3.25x10^-4^ |
|  |  |  |  |  | HSV-1 - | 0.398 | 1.74 | 0.002 |
|  | 8 | 27548680 | rs74373662 | EPHX2 | Interaction | 0.058 | 8.18 | 3.25x10^-7^ |
|  |  |  |  |  | HSV-1+ | 0.061 | 1.33 | 0.244 |
|  |  |  |  |  | HSV-1 - | 0.054 | 0.2 | 1.54x10^-5^ |
|  | 9 | 18882564 | rs1368772 | ADAMTSL1; SAXO1 | Interaction | 0.622 | 2.85 | 7.95x10^-8^ |
|  |  |  |  |  | HSV-1+ | 0.621 | 1.76 | 3.17x10^-6^ |
|  |  |  |  |  | HSV-1 - | 0.625 | 0.62 | 0.008 |
|  | 10 | 11269685 | rs61703363 | CELF2 | Interaction | 0.152 | 0.26 | 4.02x10^-7^ |
|  |  |  |  |  | HSV-1+ | 0.164 | 0.63 | 0.004 |
|  |  |  |  |  | HSV-1 - | 0.129 | 2.65 | 4.27x10^-5^ |
| EA | 14 | 23869851 | rs60016771 | LINC00596 | Interaction | 0.022 | 3.9 | 5.62x10^-8^ |
|  |  |  |  |  | HSV-1+ | 0.021 | 1.88 | 1.27x10^-4^ |
|  |  |  |  |  | HSV-1 - | 0.023 | 0.47 | 2.69x10^-4^ |
| NAH | 2 | 226385029 | rs117907276 | AC068138.1// COL4A3 | Interaction | 0.049 | 0.18 | 2.68x10^-7^ |
|  |  |  |  |  | HSV-1- | 0.058 | 3.89 | 3.13x10^-4^ |
|  |  |  |  |  | HSV1 + | NA | NA | NA |

^*^ AA = African American, AJ = Ashkenazi Jewish, CH = Caribbean Hispanic, EA/MID = admixed European and Middle Eastern, EA = European ancestry, NAH = Native American Hispanic

^†^ statistical test for the SNP in the entire sample (HSV-1 × interaction term) or in persons with (+) or without (-) HSV-1 analyzed separately

^‡^ MAF = minor allele frequency

^§^ Odds ratio

**Table 6. Genome-wide significant and suggestive (P < 5.0x10^-7^) SNP × HSV-1 interaction associations with AD risk for meta-analysis**

| **Study Group** | **Chr** | **Locus** | **SNP** | **Gene** | **P-meta** | **P-het** | **P-resid** | **Dir** |
| --- | --- | --- | --- | --- | --- | --- | --- | --- |
| ALL | 2 | 166079596 | rs115525490 | SCN1A | 4.65x10^-7^ | 7.59x10^-7^ | 0.288 | -+---- |
|  | 2 | 178128292 | rs76843678 | RBM45 | 4.42x10^-8^ | 1.11x10^-8^ | 0.076 | +++-+-- |
|  |  | 178157221 | rs11690846 | RBM45 | 4.35x10^-9^ | 1.34x10^-9^ | 0.213 | +-+-+- |
|  | 4 | 146100638 | rs12511944 | LINC01095 | 3.95x10^-7^ | 1.38x10^-7^ | 0.430 | -+---+- |
|  | 5 | 132652749 | rs139671377 | TH2LCRR | 5.39x10^-8^ | 1.43x10^-7^ | 0.029 | ++---+- |
|  | 7 | 101111535 | rs79529160 | RP11-395B7.7* | 1.35x10^-7^ | 2.31x10^-7^ | 0.934 | ---+-+ |
|  | 14 | 23862730 | rs61571833 | RP11-388E23.3 | 2.55x10^-7^ | 0.001 | 0.057 | ++-+-++ |
|  | 18 | 28344630 | rs77999338 | CDH2 | 7.52x10^-8^ | 4.94x10^-8^ | 0.753 | +---+- |
|  | 20 | 60648563 | rs1150437 | MIR646HG | 4.03x10^-7^ | 2.08x10^-7^ | 0.545 | -+-+-- |
| EU | 6 | 44196178 | rs77025261 | CAPN11//SLC29A1 | 5.56x10^-8^ | 9.32x10^-6^ | 0.187 | --+ |
|  |  | 44201302 | rs58994214 | CAPN11//SLC29A1 | 6.40x10^-8^ | 2.21x10^-5^ | 0.196 | --+ |
|  | 7 | 101111535 | rs79529160 | TRIM56//SERPINE1 | 3.01x10^-7^ | 6.45x10^-6^ | 0.042 | --- |
|  |  | 154734391 | rs55674439 | DPP6 | 2.09x10^-7^ | 0.003 | 0.347 | +++ |
|  | 9 | 112641305 | rs2995806 | KIAA1958 | 4.28x10^-7^ | 0.026 | 0.387 | +++ |
|  | 12 | 86266822 | rs78893636 | MGAT4C | 3.54x10^-9^ | 4.52x10^-6^ | 0.009 | --- |
